# Pseudo-monopolar sensing of subthalamic beta power helps to predict optimal DBS contacts in Parkinson’s Disease

**DOI:** 10.64898/2026.08.25.26361305

**Authors:** Victoria S. Witzig, Annabel van der Weide, Deborah Hubers, Bart J. Keulen, Justus Schikora, Jonathan Kaplan, Arian Memarpouri, Luisa K. Drescher, Jan Roediger, Gregor A. Brandt, Rob M.A. de Bie, P. Rick Schuurman, Martijn Beudel, Andrea A. Kühn

**Affiliations:** Department of Neurology, Movement Disorders and Neuromodulation Unit, Charité – Universitätsmedizin Berlin, Berlin, Germany; Department of Neurology, Amsterdam University Medical Center, Amsterdam Neuroscience, Meibergdreef 9, Amsterdam, the Netherlands; Department of Neurosurgery, Amsterdam University Medical Center, Amsterdam Neuroscience, Meibergdreef 9, Amsterdam, the Netherlands; Neurocure Cluster of Excellence, Charité – Universitätsmedizin Berlin, Berlin, Germany; German Center for Neurodegenerative Diseases (DZNE), Berlin, Germany

**Keywords:** deep brain stimulation, Parkinson’s disease, subthalamic nucleus, beta oscillations, local field potentials, pseudomonopolar sensing

## Abstract

**Background:** Deep brain stimulation (DBS) of the subthalamic nucleus (STN) is an effective treatment for Parkinson’s Disease (PD), but identifying optimal stimulation contacts is time-intensive. Beta-band activity (13-35 Hz) from local field potentials (LFP) correlates with motor symptoms and attenuation by dopaminergic therapy and DBS supports its role as a programming biomarker. The recently introduced Electrode Identifier (EI) feature of Medtronic Percept^TM^ neurostimulators may facilitate beta-guided contact selection.

**Objective:** To evaluate whether pseudo-monopolar STN beta power obtained using EI predicts optimal stimulation contacts and compare its performance with reconstructed bipolar recordings and MPR.

**Methods:** LFPs were recorded in 69 patients using EI and Electrode Survey (ES). Prediction accuracy was assessed using predefined ranking rules and compared with clinically selected contacts. Agreement between EI, ES, and MPR was evaluated. Motor outcome was assessed using MDS-UPDRS-III.

**Results:** EI predicted clinically selected contacts above chance (TOP1: 45%, p = 0.010; TOP2-80: 57%, p = <0.001), whereas ES exceeded chance only under more inclusive selection criteria (TOP1: 38%, p = 0.073; TOP2-80: 55%, p = 0.0021). Accuracy did not differ between methods (TOP1: p = 0.720; TOP2-80: p = 1.000). EI showed highest agreement with MPR and tended to select ventral contacts. Neither method predicted motor outcome, although EI-matched contacts showed a trend toward greater improvement. Due to technical constraints, one-third of EI recordings were excluded.

**Conclusions:** Pseudo-monopolar STN beta power provides clinically relevant information for DBS contact selection with performance comparable to bipolar approaches. Further improvements are needed before clinical implementation.

## Introduction

Deep brain stimulation (DBS) of the subthalamic nucleus (STN) is an effective therapy for advanced Parkinson’s disease (PD) [1], [2]. Identification of optimal stimulation contacts typically relies on a time-intensive monopolar review (MPR), followed by iterative parameter adjustments over several months. This process requires substantial clinical expertise and patient cooperation. Furthermore, standard MPR is usually performed in the medication OFF state depending on at least one prominent contralateral motor symptom. Therefore, programming sessions frequently last many hours, creating a substantial burden for patients and clinicians. Considering these limitations, there has been growing interest in objective and time-efficient strategies to facilitate the identification of effective stimulation contacts [3], [4], [5].

Electrophysiological studies show that beta band activity (± 13-35 Hz) in local field potentials (LFPs) recorded from DBS electrodes correlates with bradykinesia and rigidity in PD [6], [7] and is attenuated by dopaminergic medication and DBS [8], [9]. Moreover, the degree of levodopa-induced beta reduction is associated with motor improvement [6]. As beta activity is mainly localized in the dorsolateral STN [10], [11], [12], it has emerged as a promising biomarker for motor impairment and therapeutic efficacy [13]. The introduction of chronic sensing technologies, such as the Medtronic Percept^TM^ device with BrainSense^TM^ technology now enable postoperative, non-invasive LFP recordings. However, electrophysiology-guided programming [14] has not yet been widely adopted in clinical practice. To date, most clinical applications of beta-based programming rely on complex bipolar LFP comparisons between contact pairs [14], [15]. More recently, the Electrode Identifier (EI) feature, integrated into the Medtronic Percept^TM^ system, was introduced to facilitate rapid identification of contacts with high pseudo-monopolar beta-peak activity [16].

In this study, we evaluated the utility of pseudo-monopolar LFP recordings using the EI feature of Medtronic Percept^TM^ neurostimulators in a three-stage analysis. First, we investigated the ability of EI to predict clinically optimal DBS contacts. Next, we analyzed the agreement between EI-based contact selection, reconstructed bipolar methods, and MPR. Lastly, we evaluated if discrepancies between these methods were associated with motor improvements, as measured by the Movement Disorder Society Unified Parkinson’s Disease Rating Scale motor examination part (MDS-UPDRS-III) [17]. Results were compared with a reconstructed bipolar method from Electrode Survey (ES) recordings [15].

## Methods

### Participants

A total of 69 PD patients implanted with bilateral subthalamic DBS electrodes (n=138) were included (Table 1). All participants received B33005 “SenSight” directional electrodes and bidirectional Percept™ implantable pulse generators (IPGs) (Medtronic, Minneapolis, MN, USA) at Charité Universitätsmedizin Berlin and Amsterdam University Medical Center. Surgical procedures followed previously described protocols [18], [19]. Data collection in Berlin was part of routine follow-up and approved by the local ethics committees (EA2/256/20). Data collected in Amsterdam was obtained within an ongoing randomized controlled trial (ClinicalTrials.gov identifier: NCT06223399), approved by Amsterdam University Medical Center ethics committees (NL84601.018.23/NL-005038). All participants provided written informed consent, and the study was conducted in accordance with the Declaration of Helsinki.

**Table 1.** Clinical and demographic details of included patients.

| Demographics | Participants (n=69) |
| --- | --- |
| Males | 45 |
| Age at implant (years) | 64.52 ± 9.62 |
| Age at PD onset (years) | 53.75 ± 10.00 |
| Disease duration (years) | 10.77 ± 4.59 |
| LEDD pre-OP (mg/ day) | 1159.86 ± 442.16 |
| UPDRS III Med OFF pre-OP (points) | 44.57 ± 13.77 |
| UPDRS III Med ON pre-OP (points) | 22.46 ± 13.27 |

Abbreviations:
PD = Parkinson's disease
LEDD = Levodopa Equivalent Daily Dosage
OP = operative
UPDRS = Unified Parkinson's Disease Rating Scale

Each patient underwent one MPR and at least one LFP recording session including ES and EI at the first visit (one month after surgery in Amsterdam; three months in Berlin), and/or during follow-up (six months after initial programming in Amsterdam; twelve months post-surgery in Berlin). LFPs were recorded using the EI and ES functionalities of the Percept^TM^ device, and only level recordings were analyzed. Final contacts were determined during DBS optimization independently of LFP data. Motor improvement was defined as percentage change in MDS-UPDRS-III (medication OFF/stimulation OFF vs. medication OFF/stimulation ON).

### Clinical programming

In Amsterdam, initial stimulation parameters were determined using MPR during a one-day hospital admission four weeks after surgery following overnight medication withdrawal. Each level contact (four contacts per hemisphere) was systematically evaluated with stimulation amplitude increased in steps of 0.5 mA, with a fixed frequency of 130 Hz and a pulse width of 60 µs. Motor symptoms were assessed using items 3 (rigidity), 4 (finger tapping), and 17 (rest tremor amplitude) of the MDS-UPDRS-III. Side effects were recorded for each stimulation step and contact. The electrode contact providing the optimal balance between clinical benefit and side effects was selected for chronic stimulation. Stimulation parameters were optimized over six months. Follow-up was conducted during a visit six months after MPR, during which motor performance was evaluated using the MDS-UPDRS-III under all four medication and stimulation conditions (medication OFF/stimulation OFF, medication OFF/stimulation ON, medication ON/stimulation off, medication ON/stimulation ON).

In Berlin, an MPR was conducted as part of routine clinical follow up three months post-surgery following a site-specific protocol adapted from the classical MPR. For each hemisphere, one primary and one secondary target symptom was assessed using the items 3 (rigidity), 4 (finger tapping), and 17 (rest tremor amplitude) from the MDS-UPDRS-III, rated in 0.5-point increments. Baseline ratings were obtained with stimulation turned off for at least 30 minutes. Stimulation amplitude was increased in 0.5 mA steps with a fixed frequency of 130 Hz and a pulse width of 60 µs. Target symptoms were rated at each step until a first clinical effect was reached. Amplitude was further increased until saturation of benefit or side-effect threshold was identified. Consequently, the midpoint between effect and saturation/side-effect thresholds was calculated, and the mean of both symptoms defined as the testing amplitude.

At the testing amplitude, all level and segmented contacts were tested twice in randomized order, with patient and rater blinded to stimulation settings. For each contact, mean MDS-UPDRS-III scores were calculated per item for both symptoms and used to generate a contact ranking per hemisphere. Consequently, the ranking could contain two or more contacts with identical MDS-UPDRS-III scores, resulting in the possibility of multiple contacts being ranked first. Side-effect thresholds were subsequently assessed for all level and segmented contacts likewise to Amsterdam procedures. Follow-up occurred twelve months post-surgery during a subsequent hospital admission for DBS parameter optimization. If the MPR had not been performed according to the previously mentioned protocol, it was repeated. Motor performance was likewise assessed under all four medication and stimulation conditions.

### Recording procedure

Electrophysiological recordings were conducted with participants seated at rest in a medication OFF (≥ 12 hours) and stimulation-off state (≥ 30min). Each patient underwent one or two LFP recording sessions between January 2025 and March 2026: (a) during MPR one-month post-surgery in Amsterdam or three months post-surgery in Berlin, and/or (b) during follow-up six months after initial programming in Amsterdam or twelve months after initial programming in Berlin. Inclusion required the availability of one EI recording paired with an ES recording during the same visit as the MPR or during follow-up. If two recordings per patient were available, the initial recording was chosen. Raw LFP signals were sampled at 250 Hz using the standardized ES and EI functions of the Medtronic Percept™ neurostimulator (Medtronic, Minneapolis, MN, USA). Recordings were exported as JSON files.

### Statistics

For LFP data processing and all analyses, custom-written Python scripts and the following software packages were used: Python v3.13.5, NumPy v2.1.3, pandas v2.3.4, SciPy v1.15.3, Python package *specparam* (), matplotlib v3.10.0, scikit-learn v1.6.1, openpyxl v3.1.5, and IPython v8.30.0. All results are reported as mean +/- SD.

### LFP processing

#### Electrode Identifier

The El function is an automated algorithm that ranks electrode contacts based on relative LFP magnitude [20]. Recordings including levels and segments were obtained for approximately 100 seconds per hemisphere. To assess the clinical usability of the EI, we considered the default (“out-of-the-box”) EI outputs (assigned labels and LFP magnitudes) without re- calculating PSDs. LFP signals were recorded as differential signals between the selected electrode contact and the contralateral uppermost contact as reference electrode. In cases where signal amplitudes were insufficiently distinct to allow reliable ranking at a given center frequency, contacts were flagged as having “Insufficient Signal Separation”. For each channel, time-domain LFP data was transformed into the frequency domain using Fast Fourier Transform. Spectral estimates were computed using frequency bins with a width of 0.98 Hz, covering center frequencies from 0 to 96.68 Hz. Automatically processed PSDs were generated and ranked according to relative beta peak magnitude in the alpha-beta range (∼8- 35 Hz). Contacts were classified as highest (≥80% of maximum magnitude), middle (40–80%), or lowest (<40%). Rankings were visualized categorically and separately for levels and segments with three green dots, indicating the highest PSD magnitude, two grey dots indicating the middle magnitude, and one grey dot indicating the lowest magnitude. While the highest and lowest ranking was always present, multiple contacts could fall within the same ranking category (Figure 1). The algorithm initially identified a frequency at which a spectral peak was detected (selected frequency), but users could select an alternative peak frequency, upon which the contact rankings were recalculated (Figure 1).

**Figure 1.**
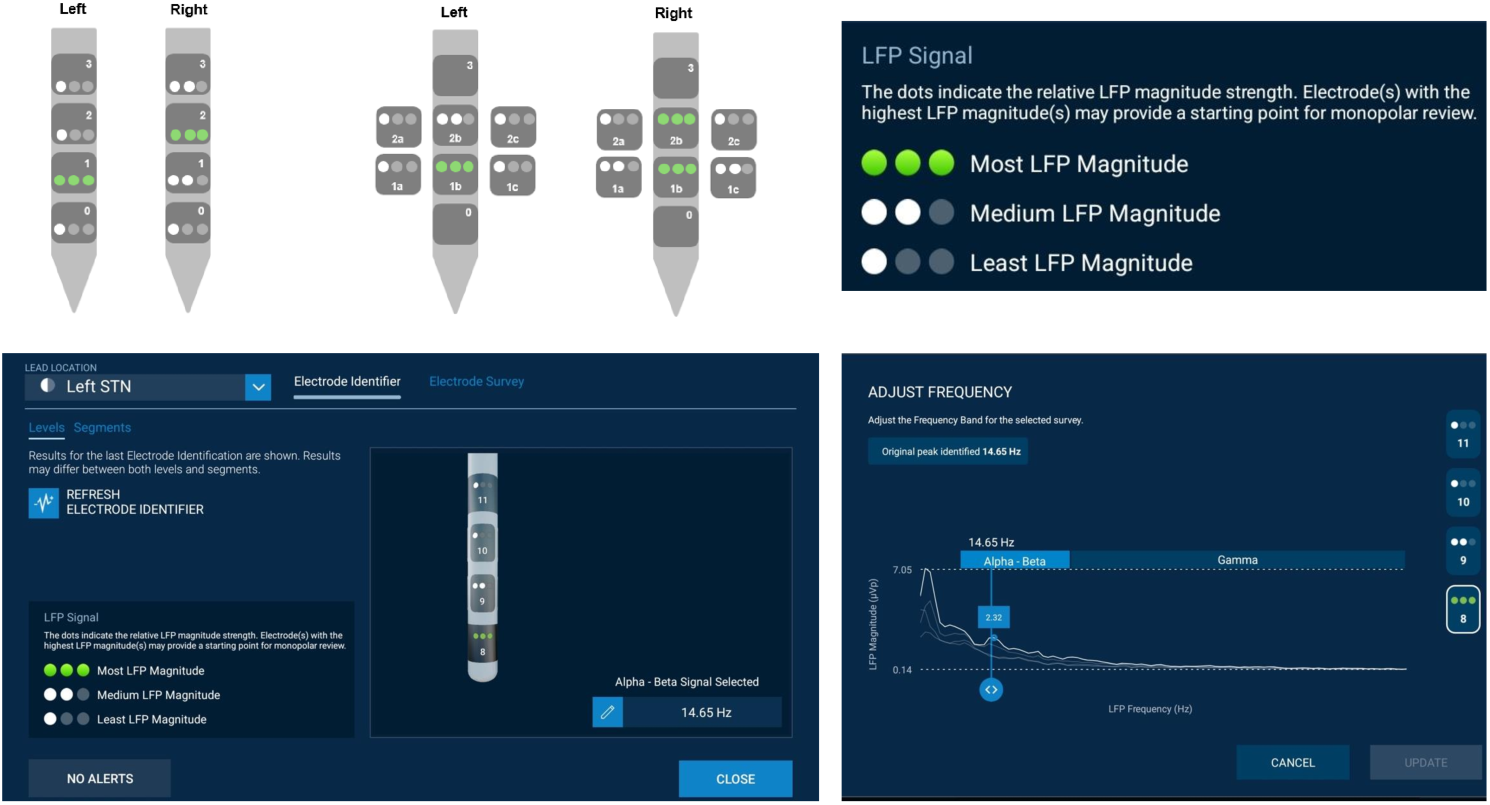
Schematic representation of Electrode Identifier ranking and Electrode Identifier user interface and advanced analysis view. Contacts are ranked according to relative LFP magnitude using dot markers. Rankings are shown per hemisphere for level and segmented contacts. Screens illustrate Electrode Identifier interface: landing screen rankings and advanced view for PSD inspection and frequency selection.

#### Electrode Survey

For each hemisphere, LFPs were recorded for 20 seconds from 15 bipolar channels per lead (six level pairs and nine segment pairs) using the ES function. Raw LFPs were visually inspected for contamination by movement artifacts and electrocardiographic activity. Components containing clear artifacts across multiple channels were removed using independent component analysis with consensus from two raters (A.W., V.S.W.). PSDs were computed using short-time Fourier transform (1-second Hanning windows, 25% overlap) and spectral beta power (13–35 Hz) averaged across the recording period. For each power spectrum, the Python package *specparam* (https://pypi.org/project/specparam/) [21] was used to fit a model over the frequency range of 2–95 Hz to isolate periodic components and identify peak center frequency and power. The parameter settings used were peak_width_limits = [3, 20], max_n_peaks = 4, min_peak_height = 0.1, and aperiodic_mode = “fixed”. Pseudo- monopolar beta power was estimated using the Euclidean distance-weighting method, which weights spectral power by the inverse squared Euclidean distance between each bipolar recording site (based on mean spatial coordinates) and the contact of interest [15], [22]. The approach incorporates all available bipolar LFP recordings (n = 15) and produces a single power spectrum for each contact, from which beta power was calculated as the average power within the 13–35 Hz range. Contacts were subsequently ranked based on their estimated beta power.

#### Prediction accuracy

Performance of EI and ES was evaluated at the hemisphere level by comparing the agreement of predicted contact sets with the clinically programmed stimulation contacts (prediction accuracy). Accuracy was computed for EI and ES under three predefined selection rules: the TOP1 rule, the TOP2-80 rule, and the label-based EI rule. Under the TOP1 rule, contacts were ranked according to their PSD score. In cases where multiple contacts shared the same value, all such contacts were included in the predicted set. Under the TOP2-80 rule, the predicted set always included the highest-scoring and the second-scoring contact if its score was at least 80% of the top-ranked score. For EI, the categorical labels provided by the system were also evaluated independently of the ranking-based rules. Under this label-based EI rule, all contacts labeled “Highest” by the EI software were selected as the predicted set. Statistical significance of prediction accuracy relative to chance was assessed using permutation-based testing for all prediction rules. For each hemisphere, method-specific ranks were randomly permuted across contacts (10,000 permutations), generating a null distribution of accuracy that preserved the number of available contacts, the within-hemisphere score distribution, and the applied decision rule (including tie handling). Permutation p-values were calculated as the proportion of permuted accuracies greater than or equal to the observed accuracy. In addition, paired differences in accuracy for TOP1 and TOP2-80 between methods were assessed using a two-sided McNemar test.

#### Agreement between methods

Agreement between EI, ES, and MPR was assessed at the hemisphere level and performed independently of the clinically selected stimulation contact at follow-up. Pairwise agreement was defined as overlap of selected contact sets (EI-ES, EI-MPR, and ES-MPR). Agreement proportions were calculated reflecting the number of hemispheres for which the relevant methods were available. Pairwise agreement matrices were generated to assess agreement between methods at the level-contact scale under the TOP1 decision rule. For each method pair (EI-ES, EI-MPR, and ES-MPR), agreement matrices were constructed by comparing the TOP1 level selected by each method, with cell counts representing the number of hemispheres. To evaluate systematic differences in level selection between methods, paired level differences were analyzed using the Wilcoxon signed-rank test. Mean signed differences and absolute differences were calculated to describe the magnitude and direction of disagreement.

#### Clinical improvement

Clinical improvement was defined as percentage change in MDS-UPDRS-III (medication OFF/stimulation OFF vs. medication OFF/stimulation ON) at follow-up. Hemispheres with ≥30% contralateral hemibody MDS-UPDRS-III improvement were classified as responders, whereas those with an improvement of <30% were classified as non-responders. For each hemisphere and prediction method, hemispheres were classified as “Match” or “No Match” based on whether the method’s predicted contact set overlapped with the final contact(s). Responder status was analyzed using population-average logistic generalized estimating equations with subject as clustering factor [23]. Continuous hemibody MDS-UPDRS-III percentage change was analyzed using linear mixed-effects models with subject-level random intercepts [24].

## Results

### Data availability

In total, 69 PD patients from Berlin (n=36) and Amsterdam (n=33) implanted with bilateral STN-DBS electrode (n=138 hemispheres) were included between January 2025 and March 2026 (Table 1). Patients had a mean age at implantation of 64.52 ± 9.62 years (mean ± SD), and the mean disease duration was 10.77 ± 4.59 years. The preoperative MDS-UPDRS-III score (medication OFF) averaged 44.57 ± 13.77 points. Pseudo-monopolar and bipolar LFPs were recorded using the EI and ES functionalities of the Percept^TM^ device, and only level recordings were analyzed. Recordings were excluded for technical reasons of the EI, including unavailable rankings, selection of frequencies outside the beta range, insufficient signal separation, or discrepancies between EI outputs and visually inspected PSDs (Figure 2). After exclusions, 78 hemispheres remained eligible for analysis. Of these, 25 lacked follow-up data on final stimulation contacts and six had segmented final contacts, leaving 47 hemispheres with corresponding final contacts (Figure 2).

**Figure 2.**
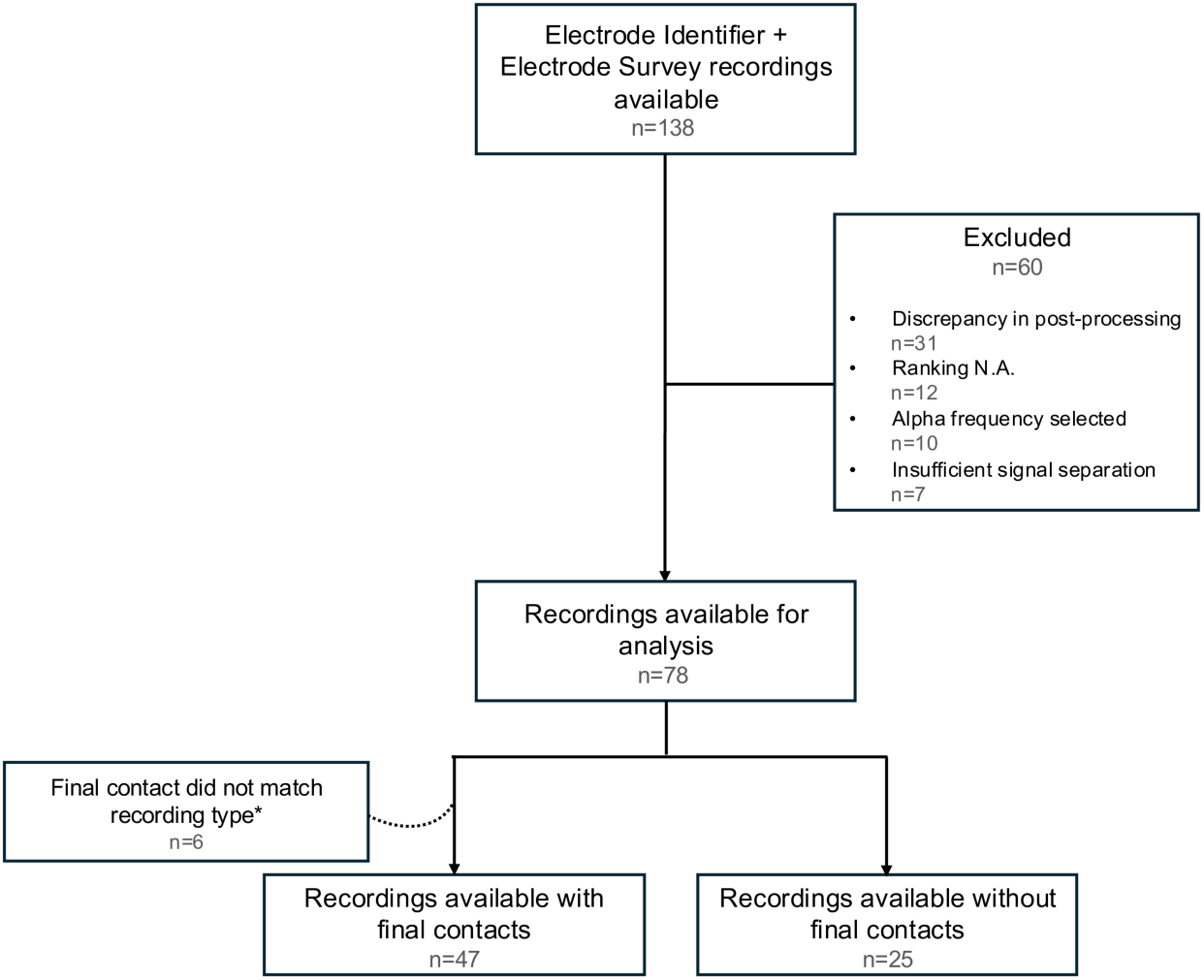
Flowchart of available recordings included in the analysis. The diagram details available hemispheres selected for analysis, indicating which level recordings were included and which were excluded at each step. Numbers represent the number of hemispheres. For each box, values are reported as n = hemispheres.

#### Prediction accuracy

Accuracy was evaluated using the above-described TOP1 rule, TOP2-80 rule, and label-based EI rule (Figure 3; Supplementary Figure 1). Using the label-based EI rule, EI achieved a mean prediction accuracy of 49%, significantly above chance (chance-level = 30%; p = 0.003). Under the TOP1 rule, EI reached 45% accuracy, also exceeding chance (chance-level = 28%; p = 0.010). In contrast, ES achieved a mean TOP1 accuracy of 38%, which did not significantly differ from chance (chance-level = 28%; p = 0.073). Despite this difference in significance relative to chance, a direct paired comparison between EI and ES using the TOP1 rule revealed no significant difference in the ability to identify clinical optimal contacts (McNemar test, p = 0.720). Naturally, application of the TOP2-80 rule increased prediction accuracies, reaching 57% for EI (chance level = 36%; p = <0.001) and 55% for ES (chance-level = 40%; p = 0.0021), with no difference between both methods (p = 1.000). To examine the impact of EI-related exclusions, we explored how ES performed in hemispheres previously removed from analysis. Across the above-mentioned exclusion categories, ES demonstrated modest prediction accuracies ranging from 36- 60% (Supplementary Table 1), comparable to its performance in the primary cohort.

**Figure 3.**
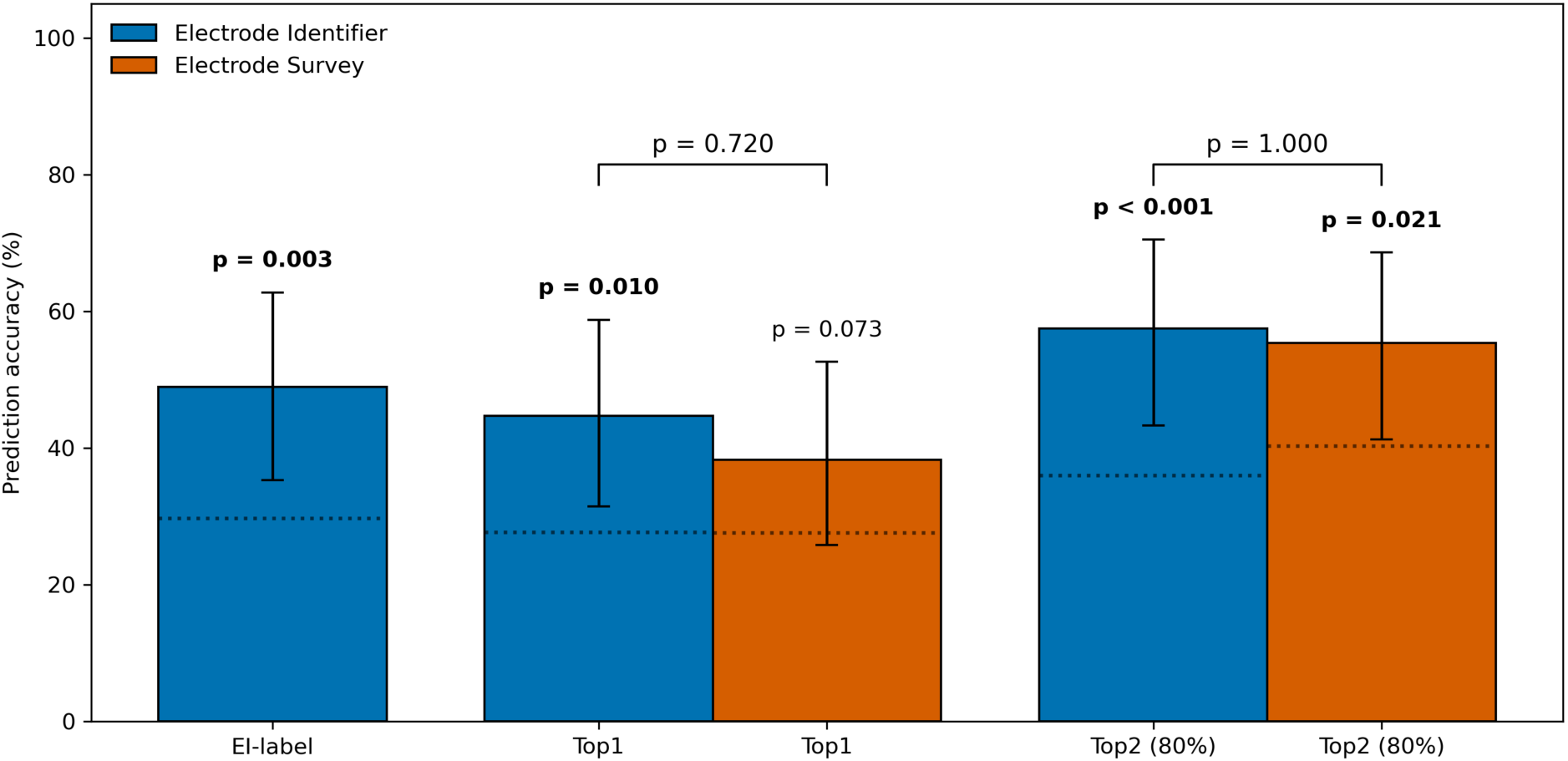
Prediction accuracy of Electrode Identifier and Electrode Survey for final contact prediction (n = 47 hemispheres) Bars indicate mean accuracy with error bars representing standard deviation and dotted line showing chance levels. Statistical significance relative to chance was assessed using permutation (p < 0.05, one-sided). Direct comparisons between methods (brackets) were evaluated using McNemar’s test (p < 0.05).

### Agreement between methods

Agreement between EI, ES, and MPR rankings was assessed pairwise (Figure 4A). Triple- agreement analyses were performed across 62 hemispheres (16 hemispheres excluded due to missing MPR results). Under the TOP1 rule, pairwise agreement was generally low to moderate. The highest concordance occurred between EI and MPR (47%), while agreement between EI and ES and between ES and MPR was lower (18% and 26%). Triple agreement across all methods under the TOP1 rule was rare (5%). When applying the TOP2-80 rule, agreement increased across all method pairs, with pairwise agreement ranging from 50-59%. Triple agreement was observed in 24% of hemispheres. Despite low overall agreement between EI and ES, selected contacts were spatially close. Disagreement between EI and ES predominantly involved adjacent contacts (mean absolute difference 0.95). In addition, ES showed a significant shift toward the dorsal segmented contact (mean signed-level difference −0.65; Wilcoxon signed-rank test, p < 0.001) (Figure 4B).

**Figure 4A.**
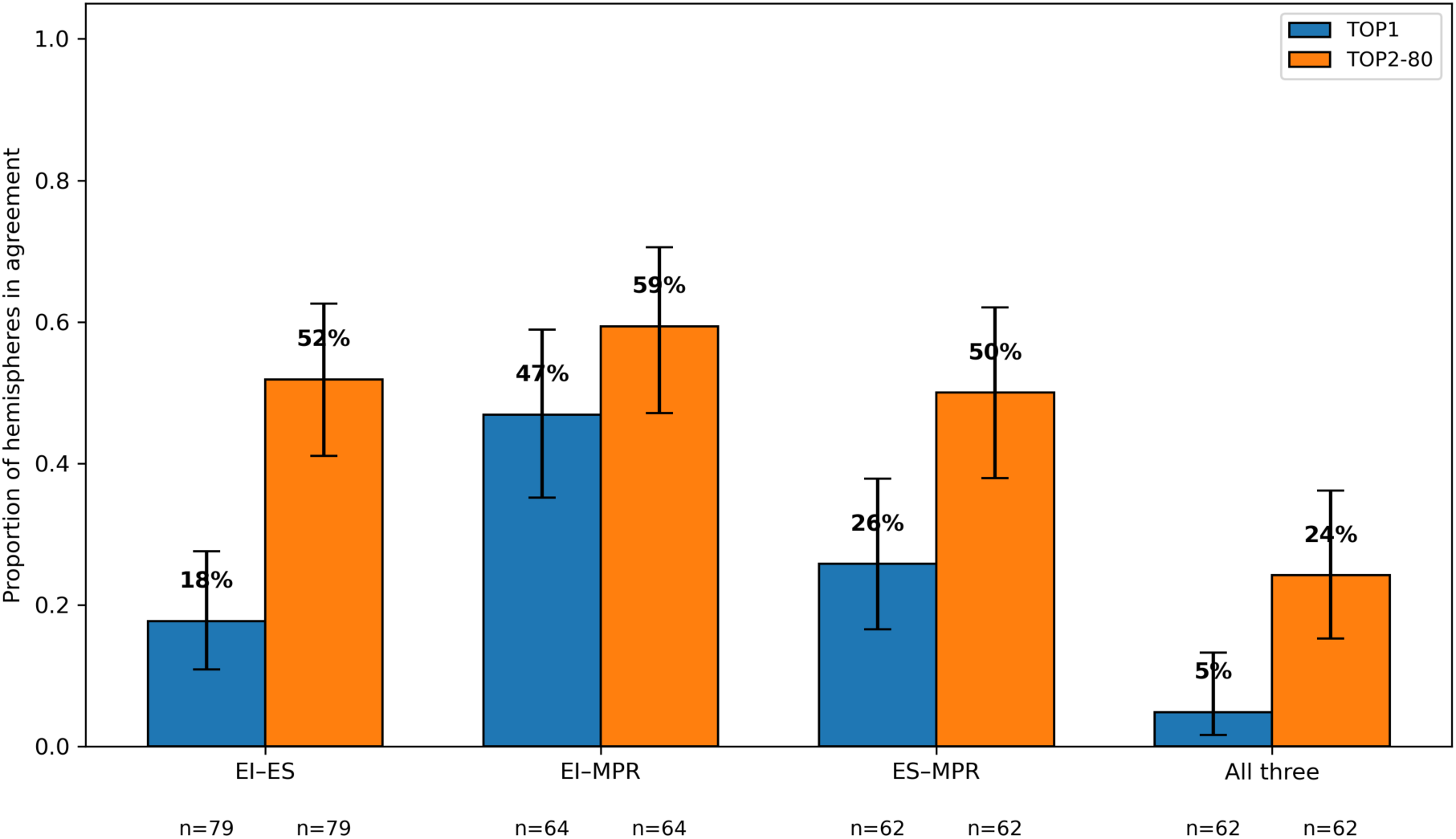
Agreement between methods. Pairwise agreement between EI, ES, and MPR for level contacts across hemispheres. Bars show proportions selecting the same contact under TOP1 or TOP2-80 rules. Error bars indicate 95% confidence intervals; sample sizes beneath corresponding bar. Hemispheres with missing data were excluded.

**Figure 4B.**
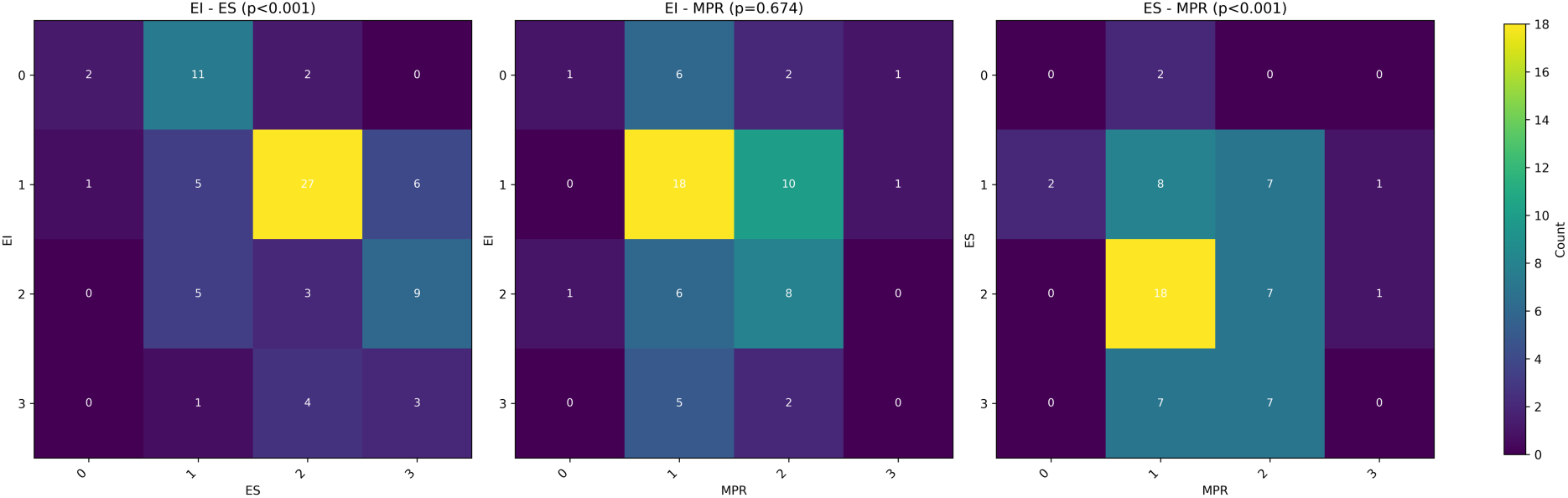
TOP1 agreement matrices. Three matrices show agreement between methods for TOP1 contact level. Cells display hemisphere counts per level pairing; color reflects counts. Data pooled across hemispheres. Paired differences were analyzed using Wilcoxon signed-rank test. Mean signed and absolute describe disagreement.

### Clinical outcome

Having an EI or ES match was not significantly associated with responder status (odds ratio = 0.71, p = 0.452 and odds ratio = 1.38, p = 0.526, respectively), and no difference between methods was observed (p = 0.448). Neither EI nor ES match was associated with hemibody MDS-UPDRS-III percentage change (β = 1.213, p = 0.813 and β = 3.97, p = 0.489), and no difference between methods was observed (p = 0.703) (Figure 5A-B). In responder-only analyses, EI match status showed a trend toward association with hemibody MDS-UPDRS-III improvement (β = 9.622, p = 0.081), whereas ES match status did not (β = 2.844, p = 0.627) (Figure 5C). Again, no difference between both methods was observed (p = 0.210).

**Figure 5A.**
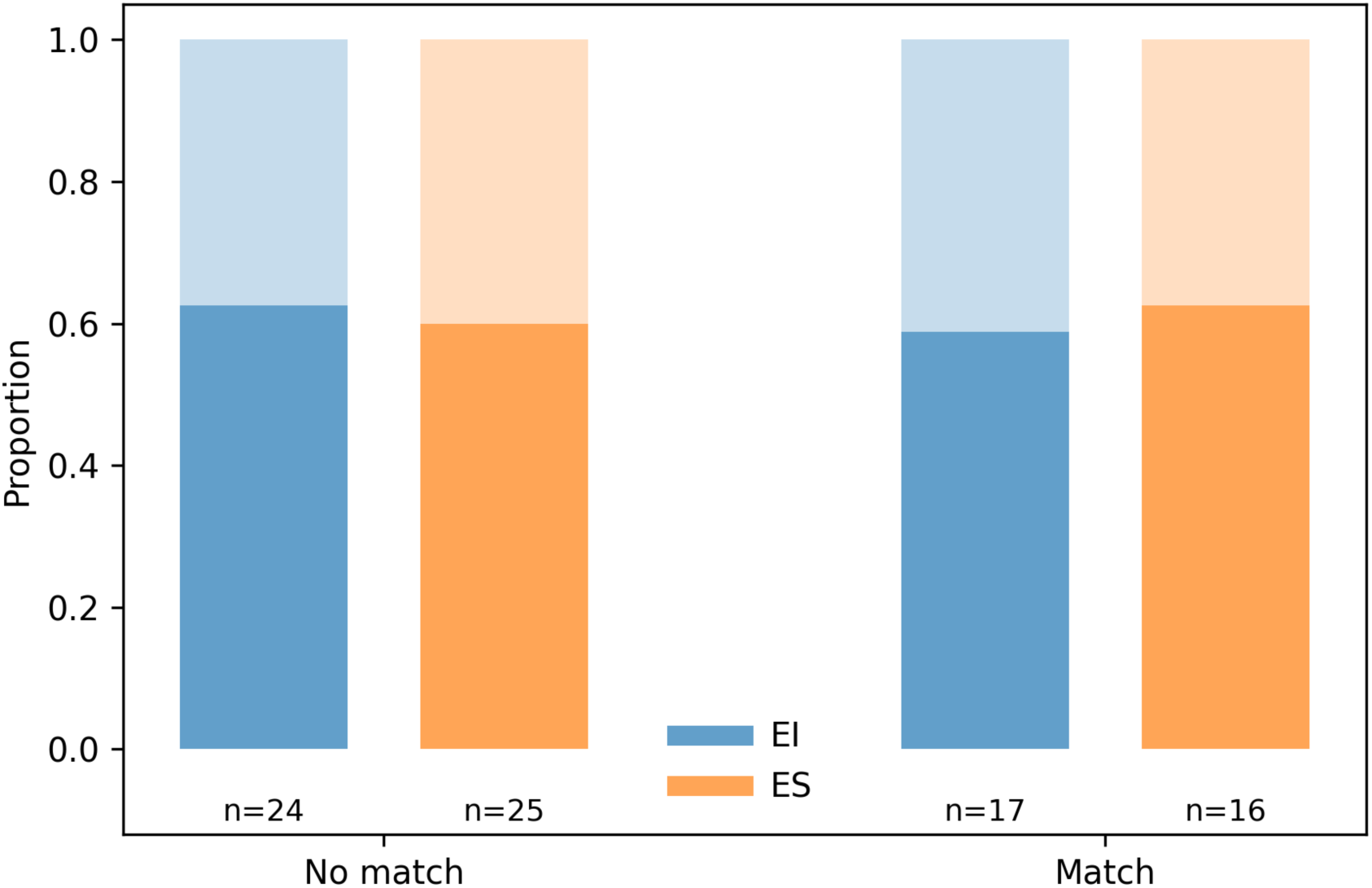
Distribution of responder hemispheres within the cohort. Proportion of responder hemispheres (≥30% hemibody UPDRS improvement) divided by match/no-match and EI/ES. Solid bar segments indicate responder hemispheres, while the transparent upper segments represent non-responders within each subgroup. Sample sizes are shown below the x-axis.

**Figure 5B.**
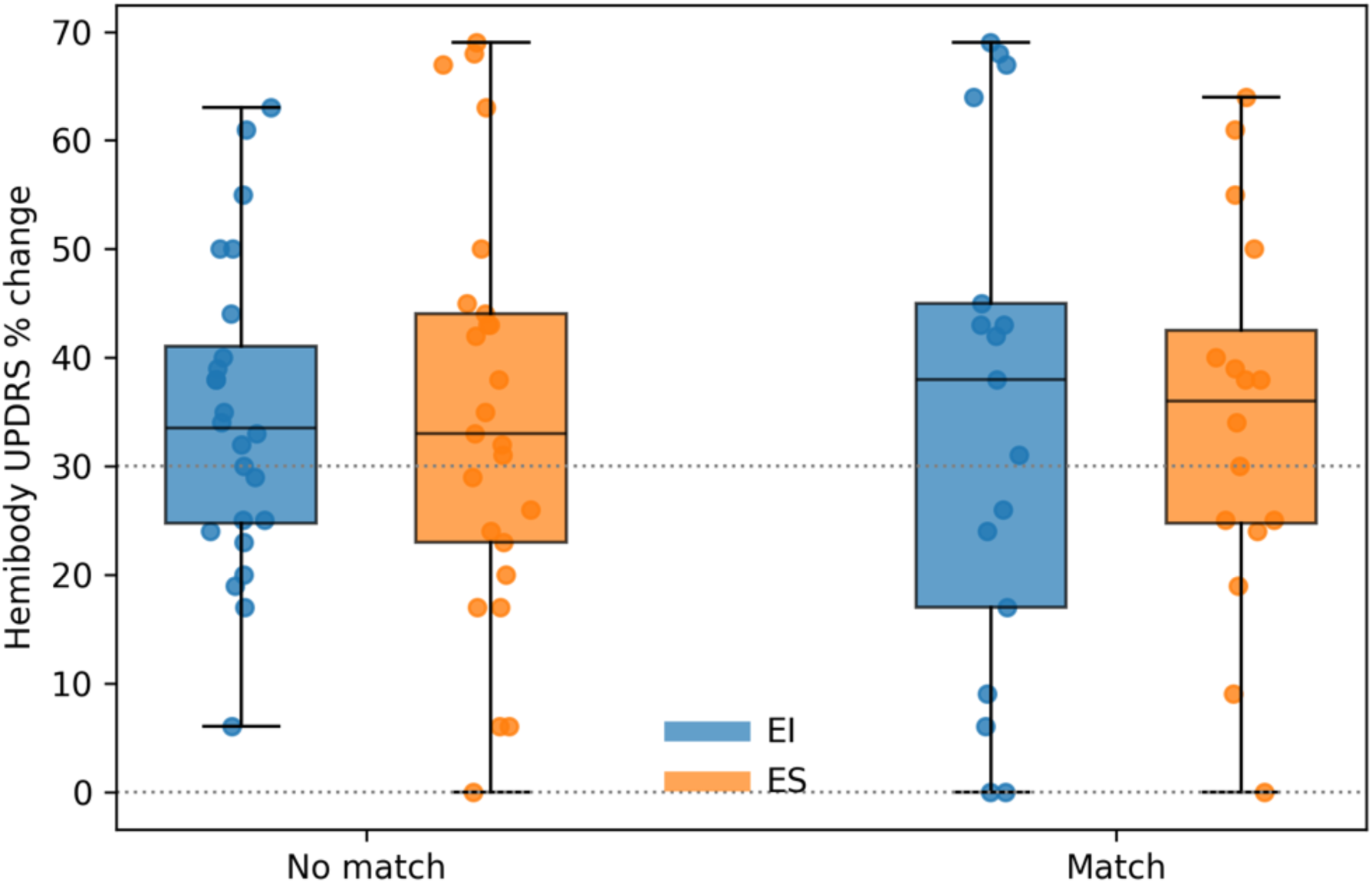
Responder status by match versus no-match status. Hemibody UPDRS-III percentage change compared to OFF-stimulation baseline across all hemispheres, divided by match/no-match and EI/ES. Single points represent individual hemispheres. The lower dotted line indicates no change, and the upper line indicates responder threshold (30%).

**Figure 5C.**
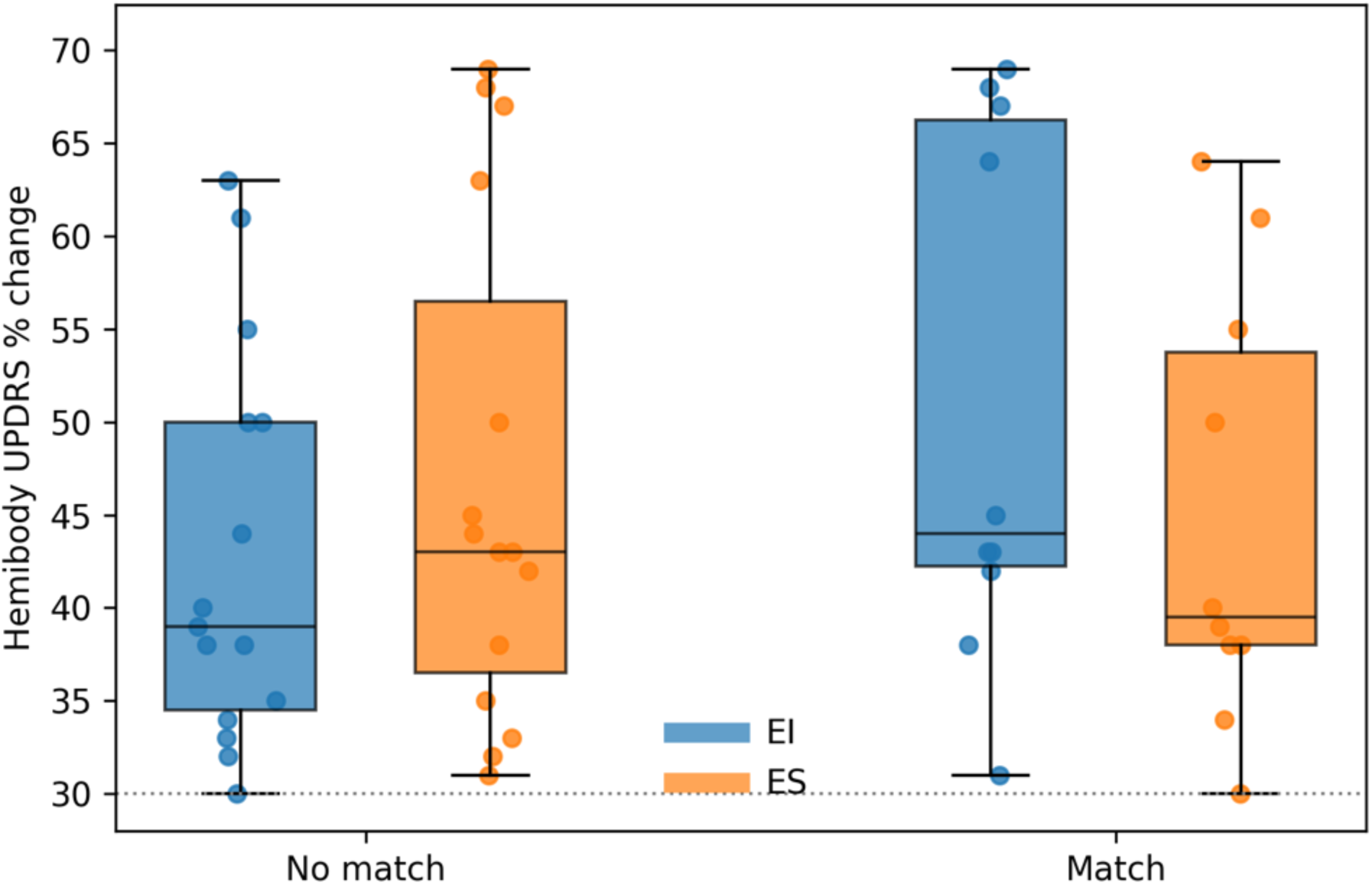
Responder-only analysis. Hemibody UPDRS-III percentage change across responder hemispheres only, divided by match/no-match and EI/ES. Boxplots indicate median and interquartile range, with individual hemispheres shown as points. Single points represent individual hemispheres, and the dotted line indicates responder threshold (30%).

## Discussion

In this study, we evaluated the clinical utility of the EI for DBS contact selection by comparing its performance with ES in predicting clinically selected stimulation contacts, assessing agreement between EI, ES, and standard MPR-based programming strategies, and investigating whether discrepancies between these methods were associated with motor improvements.

Overall, EI predicted clinically selected contacts above chance across all ranking strategies, whereas ES exceeded chance only under the TOP2-80 rule, with no difference between methods. Second, EI showed higher agreement with MPR-based clinical programming and tended to select more ventral contacts, while ES favored dorsal contacts. Third, although neither EI nor ES matches were associated with responder status or hemibody MDS-UPDRS-III improvement, hemispheres in which the clinically selected contact matched the EI prediction showed a trend toward greater motor improvement.

The comparable predictive performance of EI and ES is noteworthy given the methodological differences between the two approaches. While rankings from bipolar ES recordings were derived after extensive preprocessing, the EI was generated using semi-automated rankings by the manufacturer’s algorithm without additional post-processing. From this perspective, the comparable performance of both methods suggests that automated beta-based metrics may capture a substantial portion of the relevant neurophysiological information used to guide contact selection. While ES requires substantial analytical effort, potentially limiting its feasibility in routine clinical practice, EI can be applied rapidly with minimal user interaction.

Despite similar predictive performance, agreement analyses revealed notable differences in contact selection between methods. The probability that EI and ES would identify the same TOP1 contact was relatively low. However, closer inspection of disagreement between methods revealed a systematic shift from the dorsal segmented contact selected by ES toward the ventral segmented contact selected by EI. From a clinical perspective, this shift is relevant, as adjacent contacts are commonly explored during programming due to side effects or suboptimal stimulation outcomes [25]. On a neurophysiological level, this difference may reflect the underlying recording configurations. Bipolar sensing, as used in ES, emphasizes spatial gradients between contacts, whereas pseudo-monopolar beta estimates, as used in EI, primarily reflect the relative magnitude of local beta activity at a given contact [26]. Surgical targeting practices may further contribute to this pattern, as the most dorsal contact is often positioned outside the STN, creating beta gradients toward dorsal contacts and potentially shifting ES selections toward slightly more dorsal contacts. Consequently, ES may preferentially highlight contacts located slightly dorsal to the peak beta source, while the EI may favor contacts closer to the absolute beta maximum. Importantly, this distinction has implications for adaptive DBS. The contact suggested by EI does not necessarily correspond to the sensing configuration with the strongest beta peak in bipolar recordings. Previous work has demonstrated that optimal stimulation contacts and optimal sensing configurations may differ, reflecting the complex spatial relationship between beta generators, stimulation fields, and recording geometrics [26].

Regarding clinical outcomes, neither EI nor ES match was associated with responder status or hemibody MDS-UPDRS-III percentage improvement, and no difference was observed between both methods. However, hemispheres in which clinically selected contacts matched EI prediction, showed a trend toward greater motor improvement in responder-only analyses. Although not statistically significant, this observation suggests that pseudo-monopolar beta-based metrics may identify stimulation sites associated with greater motor benefit. If confirmed in prospective studies, these results support the fact that beta-guided pseudo-monopolar programming strategies could facilitate the identification of contacts associated with greater motor improvement.

Despite EI performing better than chance, results must be interpreted in the context of substantial data exclusion required to obtain reliable rankings. According to the manufacturer, the EI is designed as a rapid, automated tool to support initial DBS programming [20]. However, during our analysis, we found that the EI in its current “out-of-the-box” implementation did not perform robustly enough for routine clinical use. A substantial number of recordings had to be excluded due to technical constraints, which limits generalizability. EI currently requires manual inspection and adjustment of peak frequencies and therefore cannot be considered a fully automated clinical tool yet. That said, ES showed similar modest prediction accuracies in excluded hemispheres (36–60%), comparable to its performance in the primary cohort. Notably, prediction accuracy was lowest in recordings with insufficient signal separation, suggesting that limited electrophysiological signal quality may constrain contact selection irrespective of the method. These findings indicate that EI-related exclusions did not selectively remove cases in which ES performed substantially better, thereby reducing the likelihood of systematic bias in the primary analyses. An additional important limitation of this study relates to the clinical reference standard against which EI and ES were evaluated. Final stimulation contacts were defined based on routine clinical programming outcomes. However, a subset of hemispheres did not meet responder criteria on the MDS-UPDRS-III, indicating limited motor improvement under chronic stimulation. In these cases, the clinically selected contact may not necessarily reflect the therapeutically optimal stimulation site. Consequently, prediction accuracy may have been underestimated, as neurophysiology- guided methods could identify contacts associated with greater motor benefit than those selected through standard clinical programming.

In conclusion, EI shows promise as a tool supporting DBS programming. When reliable EI output was available, its predictive performance was comparable to that of ES while requiring substantially less analytical effort. However, improvements in signal handling, transparency of processing, and robustness of automated peak detection will be necessary before EI can be reliably implemented in routine clinical practice.

## Data Availability

The datasets generated and/or analyzed during this study are available from the corresponding author upon reasonable request.

---

**Supplementary Table S1.**
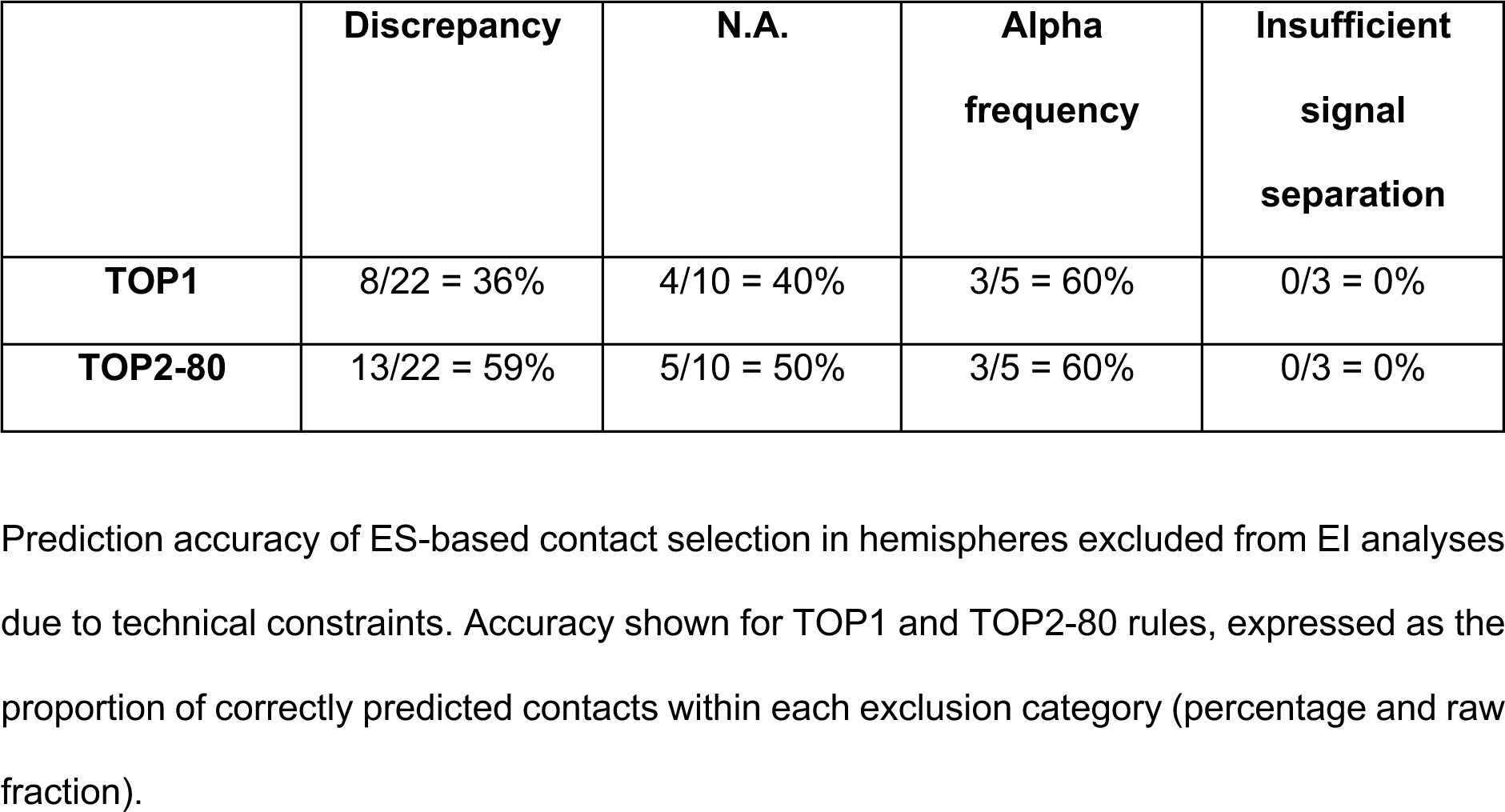
ES prediction accuracy in EI-excluded hemispheres.

**Supplementary Figure 1A.**
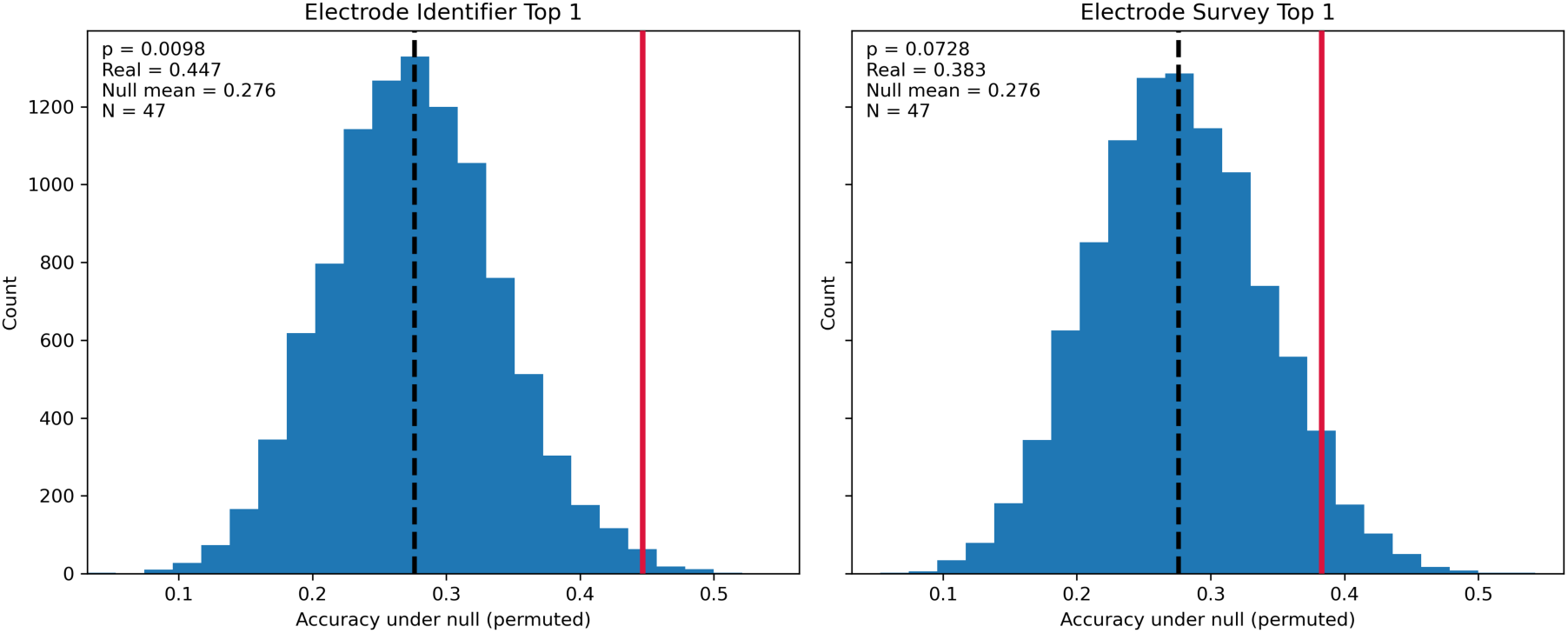
Permutation-based chance-level comparison - TOP1 method.

**Supplementary Figure 1B.**
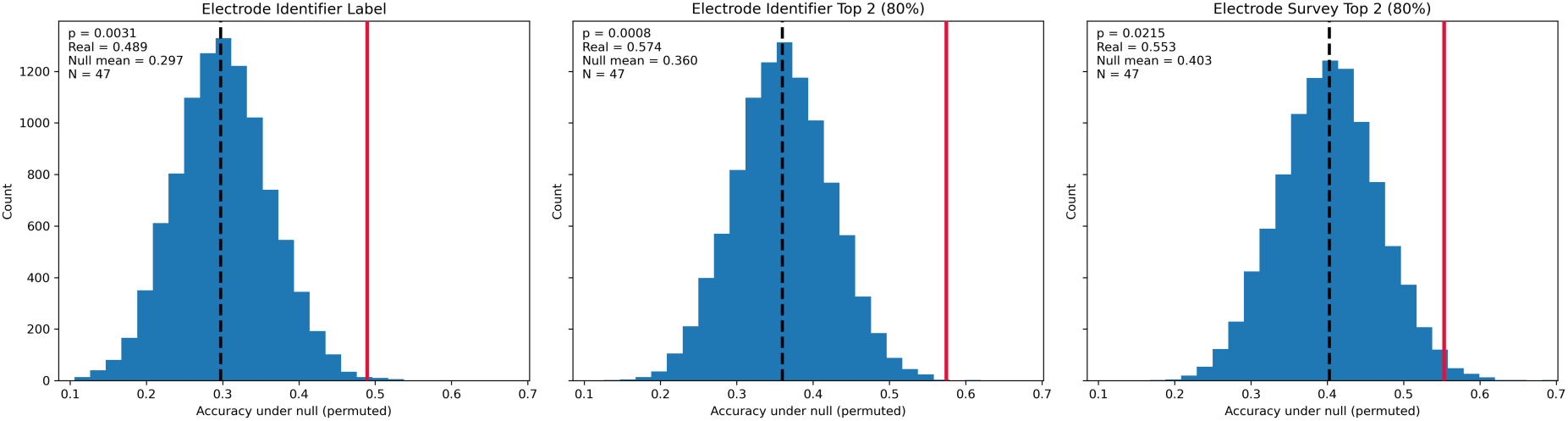
Permutation-based chance-level comparison - TOP2-80 method. Histograms show null distributions of accuracy from 10.000 permutations. Blue bars indicate permuted values, the black dashed line shows null mean, the red line indicates observed accuracy. P-values reflect proportions ≥ observed value (one-sided). N = hemispheres. Panels display EI and ES results for TOP1, TOP2-80, and EI-label.

## Author contributions

**Victoria S. Witzig:** Conceptualization, Formal analysis, Investigation, Data curation, Visualization, Writing – Original Draft, Writing – Review & Editing; **Annabel van der Weide**: Conceptualization, Formal analysis, Investigation, Data curation, Visualization, Writing – Original Draft, Writing – Review & Editing; **Deborah Hubers**: Writing - Review & Editing; **Bart J. Keulen**: Writing - Review & Editing; **Justus Schikora**: Investigation, Data curation, Writing – Review & Editing; **Jonathan Kaplan**: Investigation, Data curation, Writing – Review & Editing; **Arian Memarpouri**: Formal analysis, Investigation, Data curation, Writing – Review & Editing; **Luisa K. Drescher**: Investigation, Data curation, Writing – Review & Editing; **Jan Roediger**: Formal analysis, Methodology, Writing – Review & Editing; **Gregor A. Brandt**: Resources, Writing – Review & Editing; **Rob M. A. de Bie**: Project administration, Resources, Supervision, Writing – Review & Editing; **P. Rick Schuurman**: Resources, Writing – Review & Editing; **Martijn Beudel**: Conceptualization, Funding acquisition, Investigation, Project administration, Resources, Supervision, Writing – Review & Editing; **Andrea A. Kühn**: Conceptualization, Funding acquisition, Investigation, Project administration, Resources, Supervision, Writing – Review & Editing. All authors agreed on the final version of the manuscript.

## Acknowledgements

We would like to thank all patients who participated in this study. We would like to thank Aicha Ben Jannet for her help with organizing and assisting during recordings. We thank Jennifer K. Behnke for fruitful discussions on bipolar reconstructions.

